# One Study of COVID-19 Spreading at The United States - Brazil - Colombia^1^

**DOI:** 10.1101/2020.08.29.20184465

**Authors:** E. R. Cirilo, P. L. Natti, N. M. L. Romeiro, M.A.C. Candezano, J. M. P. Polo

## Abstract

The present work concerns the COVID-19’s spread over The United States, Brazil and Colombia. Although countries show differences in economic development, but similarities such as continental dimension or social interaction, the spread of COVID-19 in them has some similarities. At the moment, the countries are living the disease with temporal delay. Thus, we used a database on WHO Coronavirus, Mathematical Modeling and Numerical Simulations to describe the most recent COVID-19 development patterns in these countries, which we saw.

## 1. Introduction

The SARS-CoV-2 coronavirus pandemic causes the COVID-19 disease, for which we have no immune response or vaccine. The origin of COVID-19 is believed to have occurred in Wuhan, China, in late 2019. From China, the disease was rapidly transmitted globally by individuals who travelled to Europe and The United States.

On the America continent, the first cases of COVID-19 appeared in The United States. On January 21, 2020, the American Center for Disease Control and Prevention (CDC) confirmed the first case of COVID-19 in a 35-year-old man from Snohomish County, Washington, who returned from a trip to the region around Wuhan [1], In February 2020, cases of COVID-19 were already emerging in several countries on the American continent. The coronavirus landed in Latin America on February 26, when Brazil confirmed a case in São Paulo, a 61-year-old man who returned from a trip to Italy [2]. The Colombia’s first case of COVID-19 took place on March 6, 2020, a 19-year-old female who traveled to Milan, Italy [3]. Currently, COVID-19 has reached almost all countries on the American continent.

According to the World Health Organization (WHO), on July 31, 2020, The United States, Brazil and Colombia were the countries of the American continent with the highest numbers of daily infected cases [4]. By this date, 9.152.173 cases and 351.121 deaths by COVID-19 had already occurred in the American continent, comprising over 53.5% of the total number of reported cases in the world. The United States, Brazil and Colombia present 4.388.566, 2.552.265 and 276.055 cases with 150.054, 90.134 and 9.454 deaths, respectively, comprising over 48%, 28% and 3% of the total number of reported cases in the American continent [4].

The United States, Brazil and Colombia are in different moments in the epidemiological process, Colombia is in the exponential growth phase, Brazil is probably in the peak of the epidemic, while The United States is already experiencing a second wave of SARS-CoV-2 coronavirus infection. It is noteworthy that these three countries have very different Human Development Indexes (HDI) and territorial dimensions and, even so, they have in common high infection rates by the SARS-CoV-2 coronavirus.

The objective of this article is to carry out a mathematical study of the possible trends of the epidemic by COVID-19 in these three countries. This article was based on the data provided by the WHO [4] and the Susceptibles-Infectious-Recovered-Dead model, the SIRD model [5][6].

The SIRD model is a classic compartmental model of the Kermack - McKendrick type [7] [8]. Compartmental models divide the population into several different compartments, for example, Susceptible population, Infectious population, Recovered population, Dead population, among others, and specify how individuals move through the compartments over time. Despite being a simple mathematical model, the SIRD is one of the most applied mathematical models to understand the current health crisis. Obviously, more realistic and complex models would better describe the dynamics of this epidemic, but data and information about COVID-19 are lacking to implement them. Reviews of epidemiological models can be found in [5][6]

Regarding the adjustment of the parameters of the SIRD model for The United States, Brazil and Colombia, it appears that these parameters change frequently, depending on local political factors (closure of non-essential establishments, quarantine, movement restrictions…) [9], socio-economic factors (social and hygienic behaviors, lower per-capita income…) [10][11], climatic factors (temperature, humidity, average wind speed, UV index…) [12], among others. In this context, it was decided to adjust the parameters of the SIRD model over time, based on data made available by WHO, using non-linear least squares methods [13][14].

Regarding the numerical procedures to solve the SIRD model, first, the discretization of the ordinary differential equation system was performed using the Finite Difference Method. The resulting linear system of non-linear equations was solved iteratively by the Gauss-Seidel method until the convergence criterion was overcome. It was also found that all the matrices’s coefficients of the iterative processes satisfy the Sassenfeld convergence criterion [15][16] [17].

## 2. Materials and Methods

### 2.1. Data Base

There are several data base source about COVID-19, e. g., Brazil’s Coronavirus Panel [18], the National Institute of Health in Colombia [19], the Johns Hopkins Coronavirus Resource Center [20] and among other sources that exhibit the numbers of the infected, dead and more information as well.

But, to this work we used the WHO Coronavirus Disease (COVID-19) Dashboard [4] to keep integrity and uniformity of our numerical model.

From source WHO Coronavirus Dashboard we did not have the recovering data over time, so we used the assumption that says - the time for a person to be moved from the infected to the recovered compartmental can be about 14 days [21] [22]. Thus, we reconstructed the number of people in the compartments: Susceptible, Infected, Recovered and Dead.

In fact the coronavirus disease is not completely understanding at the moment, certainly there are additional parameters unknown that describing better the pattern it. So, we chose to take several data sets consolidated of the 14 days to do our simulations in order to get more realistic information. In this way, we got a simulations’ clustering which permitted to understanding the fundamentals of the behavior coronavirus disease over the countries.

How already mentioned in the introduction section, the disease begins at The United States, Brazil and Colombia countries in a different times. The government actions (good or bad), civil liberties, and mass testing come interfering with the situation of COVID-19 in countries. All of these conducts imply at the number variations of the infected and dead people, what is transcribed at the WHO Coronavirus Disease (COVID-19) data base.

From data base, it can be seen that The United States is living a second wave contamination. Brazil, maybe, arrived at the infecting peak on the August month. Colombia, clearly, is going up at the infected number yet. So, we have a time delay of the COVID-19’s spread in the countries. With these, we want to explain how disease spreading happens in the countries through numerical simulations. But with a cluster of simulations, not just a simulation alone.

### 2.2. Governing Equations

There are several mathematical modelling of the COVID-19 at the moment. But, to this work we considering the SIRD’s model given by equations:

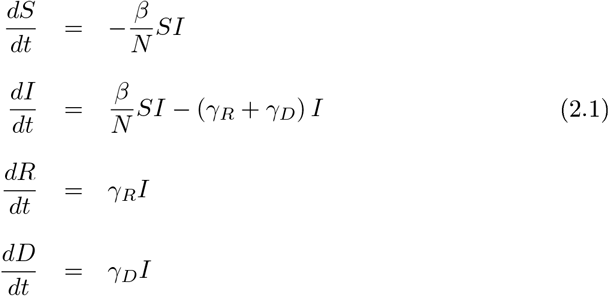

where:

*t, S, I, R, D* are time, susceptible, infected, recovered and death variables, respectively;
*N* is an average population value;
*β* is an infection rate, and *γ_R_* and *γ_D_* are recover and death rates, these parameters are calculated per day.

At the initial time we have

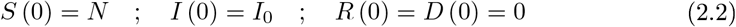

and for end time, denoted by *t_F_*, we admitted

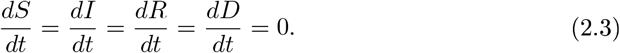

The condition (2.3) was considered because at this moment the disease finished and the equations’ steady state stays established.

### 2.3. SIRD’s Numerical Modelling Proposal

The model (2.1) has 4 ordinary differential equations (ODE). Specifically, any ODE can be written as

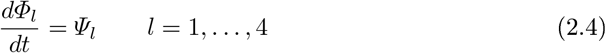

so that

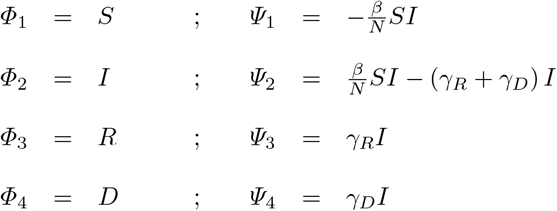

therefore, all deductive numeric procedure at this work will be over equation (2.4).

The temporal discret grid considered is showed at the Figure (1). We denoted the time lapse like Δ*t*, and the end time *t_F_* = (*s* − 1) Δ*t*. The value *k* is a time counter, thereby *k* = *s* means temporal nodes’ total value.

**Figure 1:**
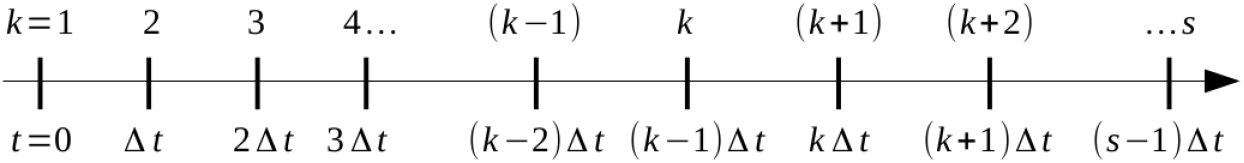
Temporal discret grid.

From condition (2.2) we can write 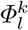 like that

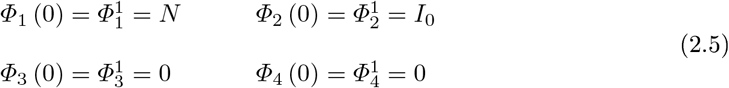

and the Neumann condition (2.3) we have too

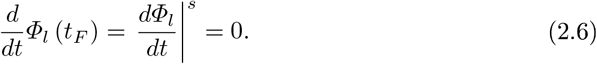

With respect to temporal derivative of the equations (2.4), they are approximated by forward second order finite difference in the node *k*

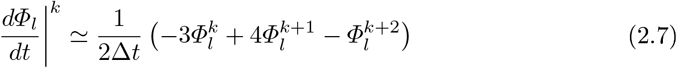

similarly, the equations (2.6) are approximated by backward second order finite difference at the last node *s*

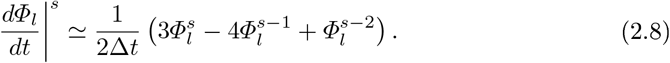

These approximations were used to obtain improved numerical results.

So, writing equation (2.4) to node *k* we can see

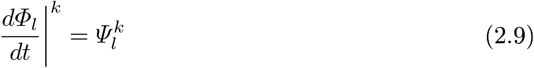

and inserting formula (2.7) at the last equation (2.9), what it leads to

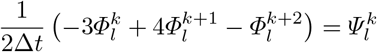

or finally

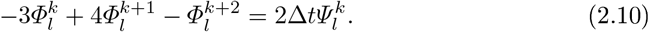

The equality (2.10) is an our temporal difference equation, that describe the COVID-19 behaviour. We want to explain that the equation (2.10) is the temporal evolution for *Φ_l_*. Numerically, from Figure (2), setting *s* we have to vary *k* = 1,…, *s* − 2 to obtain the differences’ equations at the points *k* = 2,…, *s* − 1.

**Figure 2:**
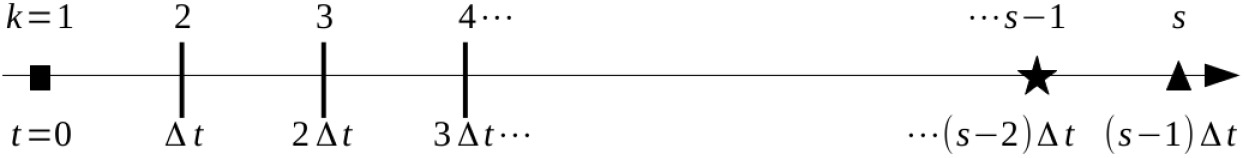
Computational temporal grid until *s*.

The first possible value is *s* = 3, so just a simple linear equation is found, and the 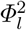 is known at the time *t_F_* = Δ*t*.

Nevertheless, with help the Figure (2) again, if *s* > 3 we will have to solve *l* systems of the (*s* − 2) difference equations to compute 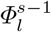 (labeled to star symbol) to *t_F_* = (*s* − 2) Δ*t*. It remembering which at the square symbol the initial condition (2.5) is assumed and the triangle symbol the Neumann condition (2.6) is used as well. Particularly, the system mentioned is the kind of

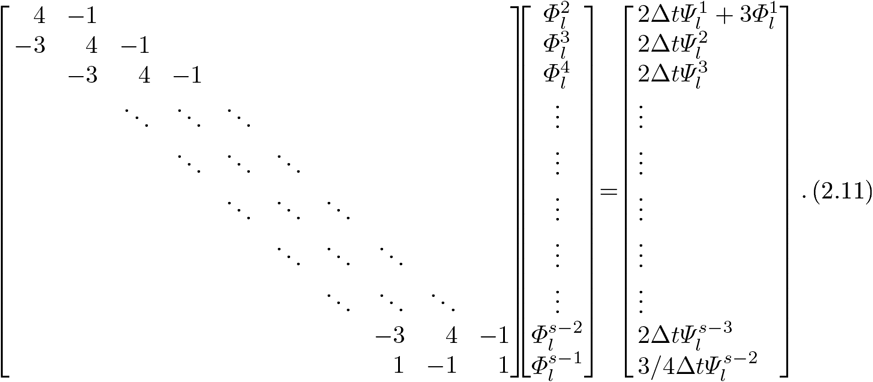

Note that the system (2.11) is not a linear system. The terms

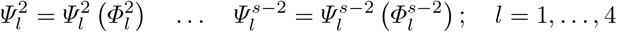

are known if the 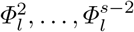 values were explicit. But they are unknown! Thus, we use a iterative strategy to solve the system (2.11) how presented in [23].

Without loss of generality, if *s* = 4 the system (2.11) becomes

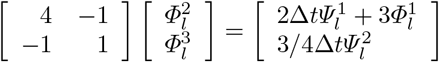

whose the iterative process associated can be like

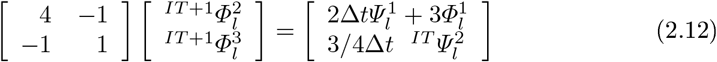

where *IT* = 0, 1,…, *IT_MAX_*, and *IT_MAX_* means iteration maximum number. Particularly, we have a linear system to each specific *IT*. Furthermore, we choose the Gauss-Seidel method to solve (2.12) because the convergence is guaranteed, it is easy to see that the Sassenfeld criterium is verified. The stopped criteria value of the Gauss-Seidel method used in this work was 10^-7^.

When IT = 0 the system (2.12) becomes

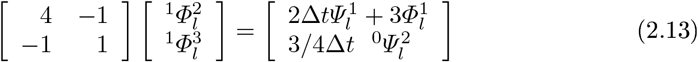

so that:

- 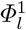 is known (initial condition);
- 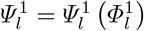 is known, of course;
- 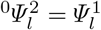 is admitted how our prediction hypothesis.

Thus, we have the linear system’s solution 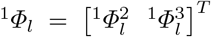 by means of Gauss-Seidel method. Now, if *IT* = 1 we find

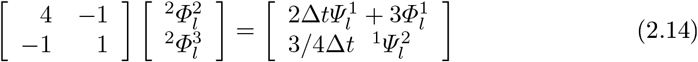

so that 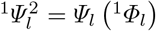 is our prediction hypothesis again. It solving (2.14), we get 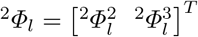.

After this, if the ^2^*Φ*_2_ is less than one (which means an end to the spread of covid-19) we accept the ^2^*Φ_l_ ≡ Φ_l_* as numerical solution of the SIRD’s (2.4) at the time *t* = 2Δ*t*. Otherwise, we set *s* = 5 and do the all process again to a new time *t* = 3Δ*t*, and so on.

The numerical process finishes if we get the *^m^Φ_2_* < 1 to some 0 *< m < IT_MAX_*. Finally, we have some gains with strategy provided here:

- the governing equations system (2.4) is solved by a convergent methodology to any time;
- our implicit methodology does not need a severe restriction on the Δ*t* value.

The implicit method together with Gauss-Seidel and Lax’s law guarantee it. Besides, the *t_F_* time is discovered in the computational run’s time. To performance all simulation we developed a fortran 90 code that solve the SIRD’s governing equations from auxiliary conditions.

### 2.4. Parameters’ Optimization

The parameters’ optimization in the SIRD (2.1) is done as in [24, 25], that is, solving a nonlinear least square problem. We define the vector function **u**(*t*) = (*S*(*t*)*, I*(*t*), *R*(*t*)*, D*(*t*)), the vectors of parameters **q** = (*β,γ_R_*, *γ_D_*) and the known data **y** at times *t*_1_,..*.t_n_*. Given a function *F*(**u**, **q**), we estimate the parameters **q** by solving the following nonlinear least square problem

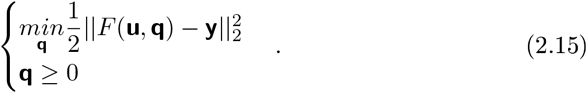

The trust-region based method adapted in the Isqnonlin Matlab function is used for the constrained optimization problem (2.15).

## 3. Discussion and concluding remarks

In this section we introduce our analyse of the COVID-19 and simulations as well. The reader will see here that the country’s richness, its continental dimension or social interaction are not avoid the Sars-CoV-2 infection.

By other hand, it is good to explain we used normalized measures, what allow us to compare the disease spread at the countries. We setting N population values like 328.200.000, 211.000.000, 49.650.000 to The United States (USA), Brazil (BRA) and Colombia (COL), respectively. Thus the SIRD’s model variables kept at the [0,1] window. Also, the times counting begins in *t* = 0 which corresponds to 2020-01-20 (USA), 2020-02-26 (BRA) and 2020-03-06 (COL). We remember that our simulations were done with data taken every 14 days to each country. What it led to label the simulations like *S*_1_,…, *S*_6_.

The Figures (3)-(4) show the data for susceptible, infected, recovered and dead in the time. We preferred to display them this shape to see the COVID-19 rises at the countries. For latin countries, the infected number came going up, but it looks like the Brazil already reached the peak. The american population is living a second wave and walks to second peak, probably. Additionally, the recover and dead people increase. But with a recover number much higher than the death.

**Figure 3:**
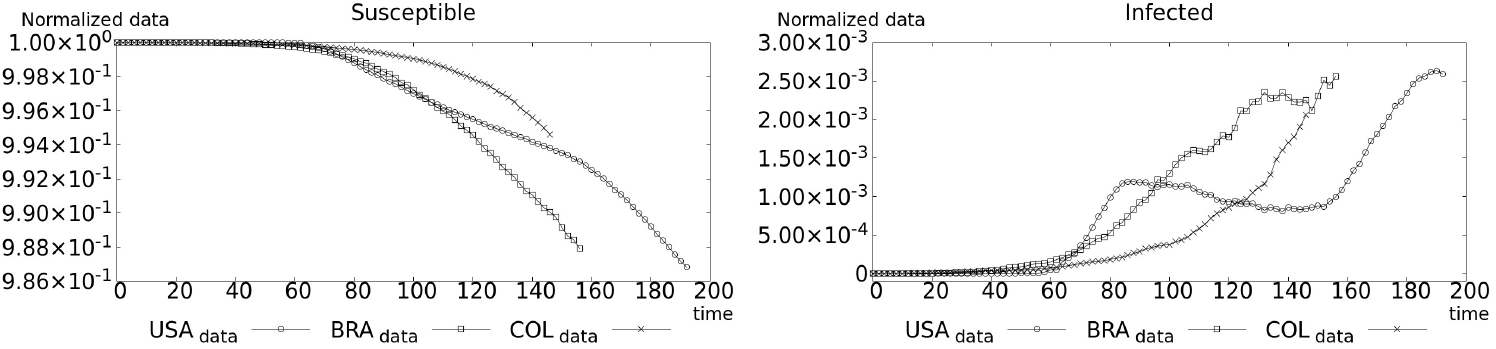
Susceptible (left), and infected (right) normalized data

**Figure 4:**
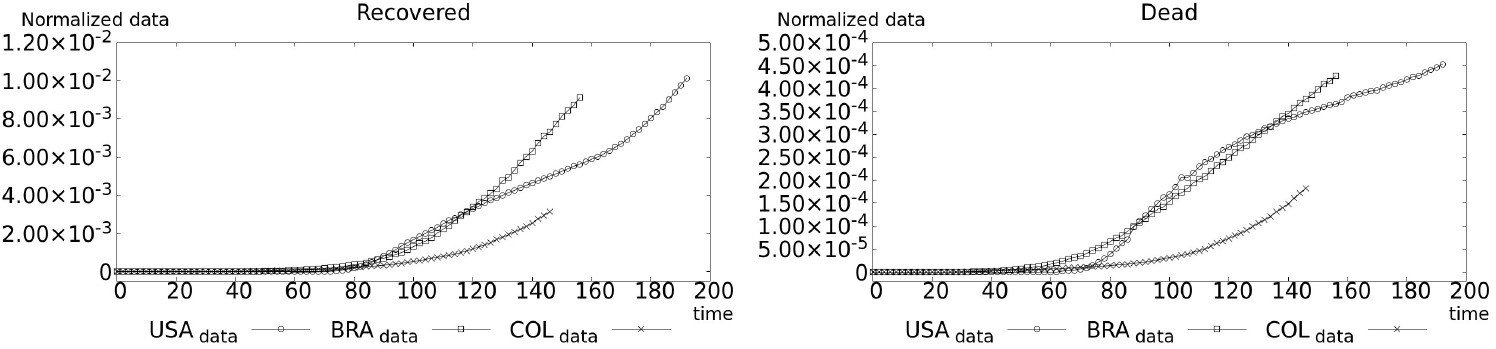
Recovered (left), and dead (right) normalized data

The Figure (5 - left) shows the susceptible decrease dynamic of The United States, and our simulations. We started simulations at the date 2020-05-04. From Figure (5 - right), the zoom Figure (5 - left), it is possible to see a changing of the disease’s drop behavior near of *t* = 139 (date 2020-06-07) by our simulations. Here, in the neighborhood of the date, it seems that there is an inflection point. This is more evident, if we look at the Figure (6) as well.

**Figure 5:**
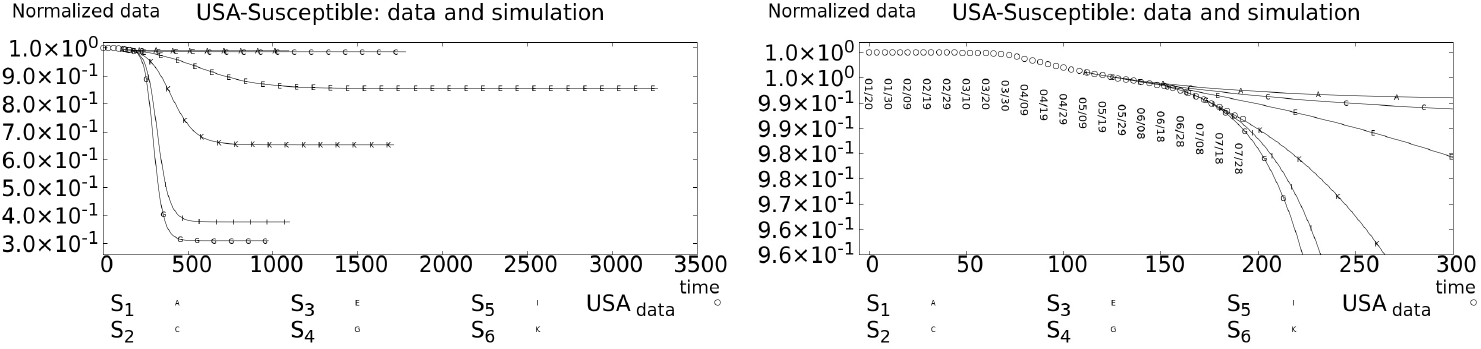
USA-Susceptible reconstructed data from WHO Coronavirus Disease (COVID-19) Dashboard [4] and simulations *S*_1_,…,*S*_6_.

**Figure 6:**
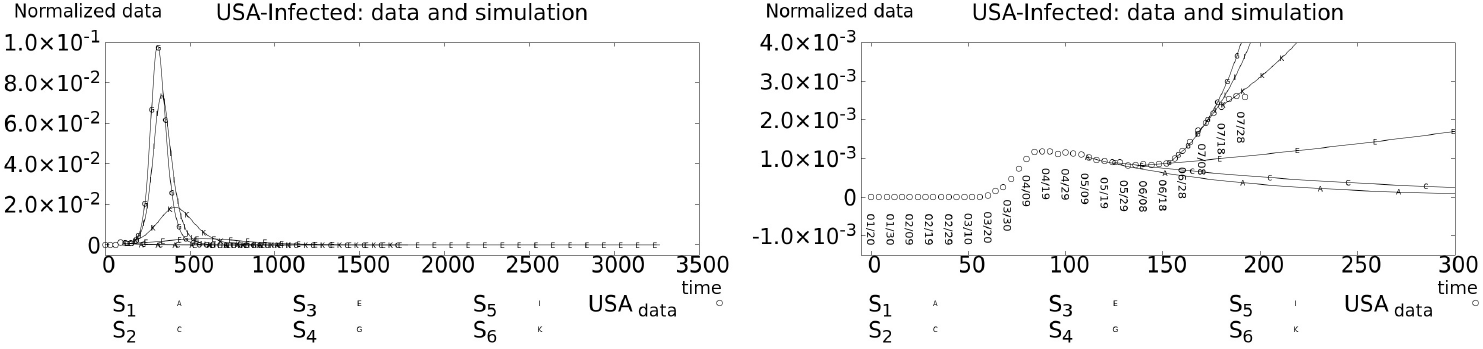
USA-Infected reconstructed data from WHO Coronavirus Disease (COVID-19) Dashboard [4] and simulations *S*_1_,…,*S*_6_.

We emphasize that the date 2020-06-07 corresponds to a 14 days after the George Floyd’s assassinate (2020-05-25) at the Minneapolis-US. Maybe, the inflection point could be correlated to not social distancing of the USA people manifestation about the assassinate. Which could to evidence the start of the second wave of Sars-CoV-2 infection.

The Figure (6 - right) shows the inverse tendency over infected people. The COVID-19 changes from deceleration to acceleration status. For we this is a characteristic of a new infection wave. The simulations predicts a new peak arising at the country, which can be seen at the Figure (6 - left). However, it is seen that the peak is being attenuating at the country.

Beyond of that, The United States have indicated a value climb at the recover and death’s compartments. But, we emphasized the recovered number’s magnitude is higher than the death number, see the Figure (7). Particularly, by our last simulations, it can be seen a damping of the accumulated numbers as well.

**Figure 7:**
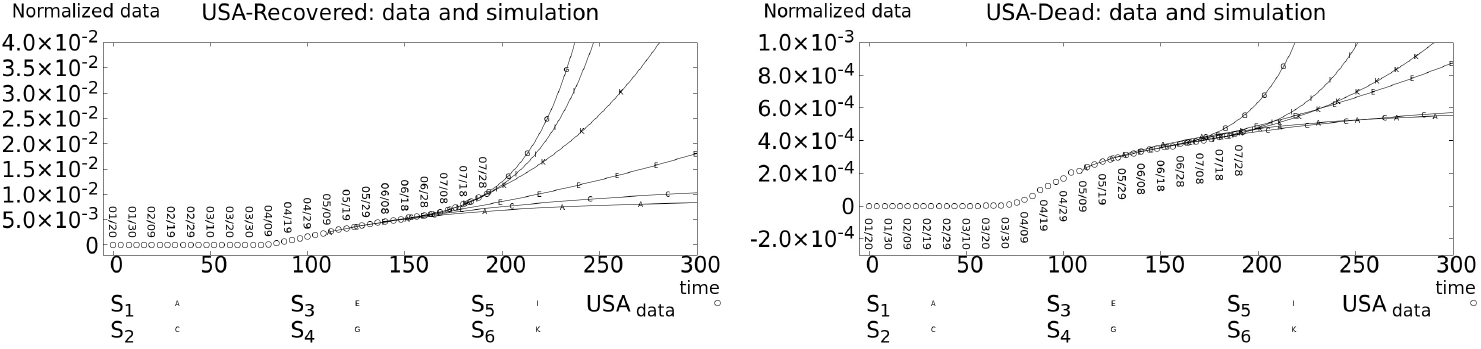
USA-Recovered(left)/Dead(right) reconstructed data from WHO Coronavirus Disease (COVID-19) Dashboard [4] and our simulations *S*_1_,…*,S*_6_.

For Brazil, the Figure (8) shows the susceptibles’ decelerating dynamic, and our simulations too. The our simulations started in the date 2020-05-12 because the COVID-19 cases became more manifest. Different of The United States, by means of our predictions, Brazil and Colombia do not live a second infection wave so far. Colombia, whose simulations started in the date 2020-05-14, presents the same patter of Brazil’s susceptible, see the Figure (9). However, the Colombia’s Covid-19 disease is more accelerated than Brazil, it’s further evident in the last days, see the Figure (8 - left) and Figure (9 - right).

**Figure 8:**
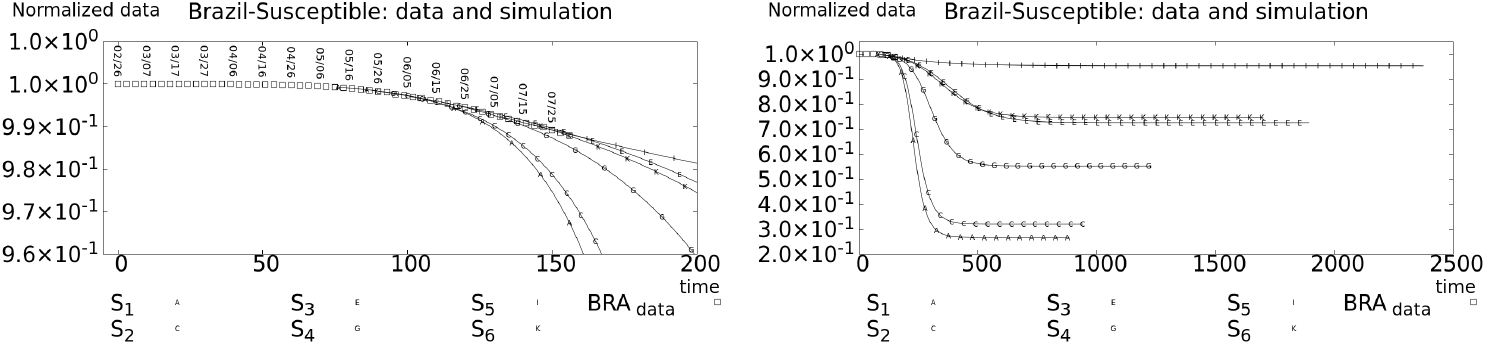
Brazil-Susceptible reconstructed data from WHO Coronavirus Disease (COVID-19) Dashboard [4] and simulations *S*_1_,…*,S*_6_.

**Figure 9:**
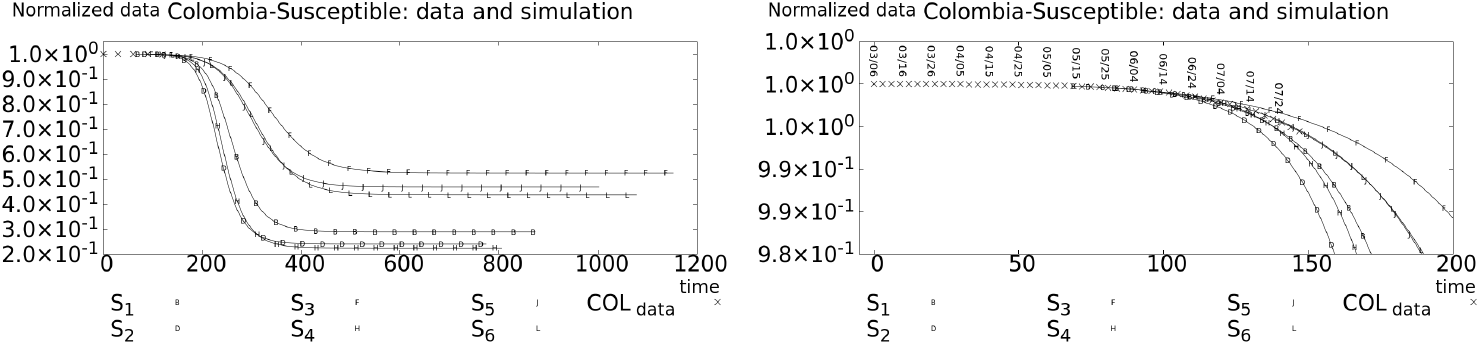
Colombia-Susceptible reconstructed data from WHO Coronavirus Disease (COVID-19) Dashboard [4] and simulations *S*_1_,…*,S*_6_.

Colombia do not rich the disease peak, yet. This can be seen through of the Figure (10). The COVID-19’s spread come increasing and its peak is still far away. Nevertheless, to Brazil it is possible to infer the disease peak and plateau happen between August and September in the present year, see the Figure (11). We assume it because at the months May and June the COVID-19 became decelerating.

**Figure 10:**
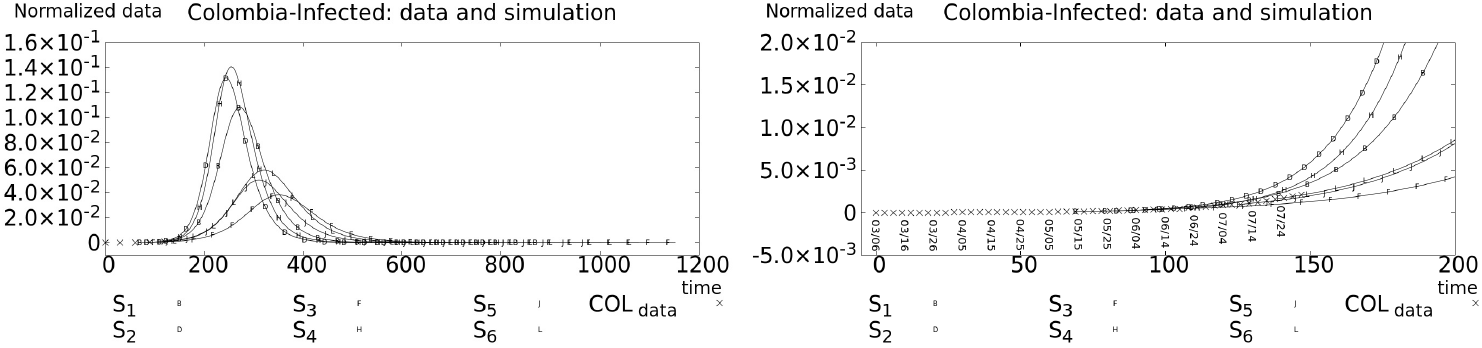
Colombia-Infected reconstructed data from WHO Coronavirus Disease (COVID-19) Dashboard [4] and simulations *S*_1_,…*,S*_6_.

**Figure 11:**
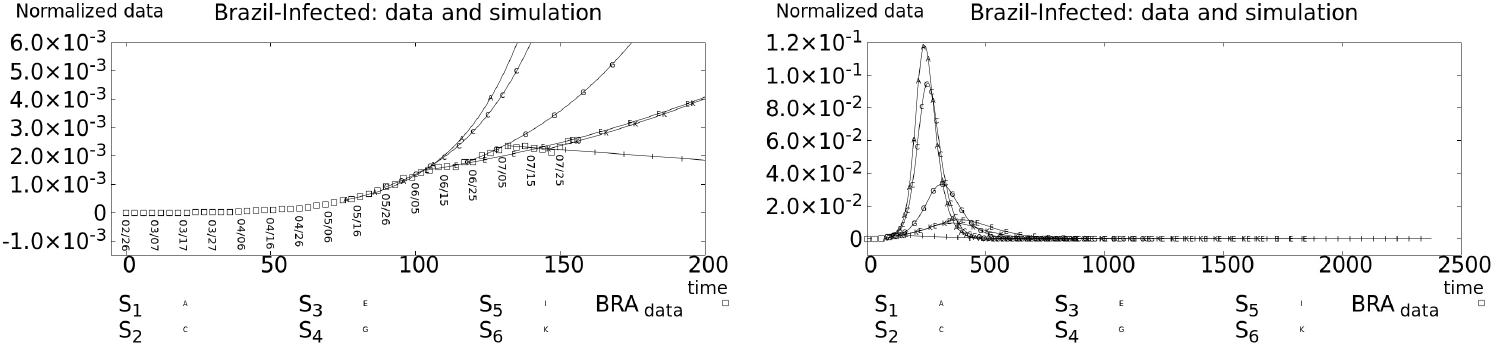
Brazil-Infected reconstructed data from WHO Coronavirus Disease (COVID-19) Dashboard [4] and simulations *S*_1_,…*,S*_6_.

Analogous to The United States, the recovered and death Brazilians numbers climbs at the values, Figure (12) but with different magnitudes between them as well. This patter happens to Colombia as well, look the Figure (13).

**Figure 12:**
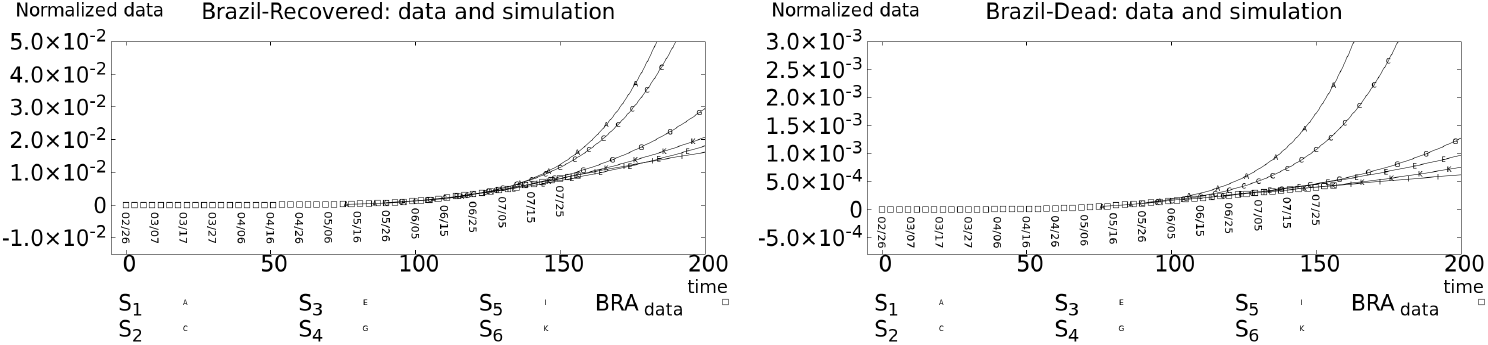
Brazil-Recovered(left)/Dead(right) reconstructed datafrom WHO Coronavirus Disease (COVID-19) Dashboard [4] and our simulations *S*_1_,…*,S*_6_.

**Figure 13:**
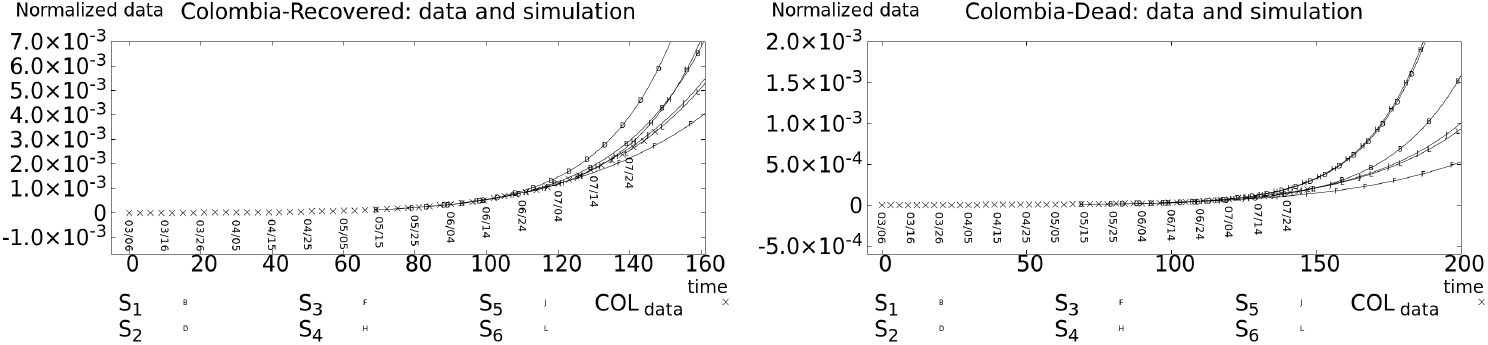
Colombia-Recovered(left)/Dead(right) reconstructed data from WHO Coronavirus Disease (COVID-19) Dashboard [4] and our simulations *S*_1_,…*,S*_6_.

We calculated the basic reproduction number 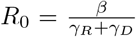 for the last simulations on July. The United States is fighting with disease, its *R*_0_ comes decreasing, see the Figure (14 - left). The Brazil’s *R*_0_ came dropping but it changed, Figure (14 - center). If it persists, it’s possible increasing infection again. The Colombia’s *R*_0_ is oscillating, Figure (14 - right), and all care are necessary at the currenty time.

**Figure 14:**
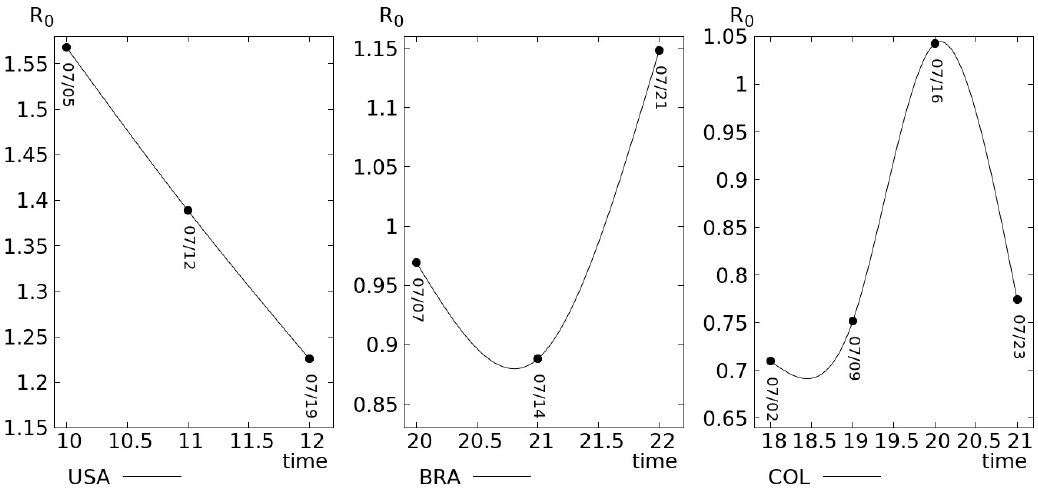
Basic reproduction number *R*_0_ at the time.

Finally, we emphasize that the COVID-19’s dynamic of the Brazil and Colombia are the same, but as delay. Of course the next days are crucials, the countries cannot relax your fight actions against COVID-19. The our simulations predict that there are some control over COVID-19 disease in theses countries, yet. Clearly, we do not know the specific proceedings which provide it. But if they relax, the countries could have a second wave infection similar to the one the United States is currently experiencing.

> **Resumo**. O presente trabalho trata do espalhamento da COVID-19 sobre os Estados Unidos, Brasil e Colômbia. Embora os países exibam diferençias no desenvolvimento econômico, mas similaridades tais como dimensão continental ou interação social, a propagação do COVID-19 neles tem algumas semelhanças. Agora, os países estão vivendo a doença com atraso temporal. Assim, usamos um banco de dados de Coronavirus da OMS, Modelagem Matemática e Simulações Numéricas para descrever o padrão mais recente de desenvolvimento do COVID-19 nesses países, que nós observamos.
>
> **Palavras-chave**. Modelagem Matemática, Simulações Numéricas, COVID-19.

## Data Availability

The data are available from.

## References

[1] L. Sun, “First u.s. case of potentially deadly Chinese coronavirus confirmed in Washington state. the Washington post.” https://www.washingtonpost.com/health/2020/01/21/coronavirus-us-case/, January 2020.

[2] A. Rodriguez-Morales, V. Gallego, and e. a. Escalera-Antezana, J.P., “Covid-19 in latin america: The implications of the first confirmed case in brazil,” Travel Med. Infect. Dis., vol. 35, p. 101613, 2020.

[3] “Colombia confirma su primer caso de covid-19. boletín de prensa, 050.” www.minsalud.gov.co/Paginas/Colombia-confirma-su-primer-caso-de-COVID-19.aspx, 2020.

[4] W. H. Organization, https://covidl9.who.int/.

[5] F. Brauer, C. Castillo-Chavez, and Z. Feng, Mathematical Models in Epidemiology. Springer-Verlag New York, 2019.

[6] Z. Ma and J. Li, Dynamical Modelinq and Analysis of Epidemics. WORLD SCIENTIFIC, 2009.

[7] W. Kermack, A. G. McKendrick, and G. T. Walker, “A contribution to the mathematical theory of epidemics, part i,” Proceedings of the Royal Society of London. Series A, vol. 115, pp. 700-721, August 1927.

[8] W. Kermack, A. G. McKendrick, and G. T. Walker, “Contributions to the mathematical theory of epidemics, ii - the problem of endemicity.,” Proceedings of the Royal Society of London. Series A, vol. 138, pp. 55-83, October 1932.

[9] D. Jorge, M. S. Rodrigues, M. Silva, L. Cardim, N. da Silva, I. Silveira, V. Silva, F. Pereira, S. Pinho, R. Andrade, P. P. Ramos, and J. Oliveira, “Assessing the nationwide impact of covid-19 mitigation policies on the transmission rate of sars-cov-2 in brazil,” medRxiv, 2020.

[10] D. Chu and et al., “Physical distancing, face masks, and eye protection to prevent person-to-person transmission of sars-cov-2 and covid-19: a systematic review and meta-analysis,” The Lancet, vol. 395, pp. 1973-1987, June 2020.

[11] W. de Souza and et al., “Epidemiological and clinical characteristics of the covid-19 epidemic in brazil,” Nature Human Behaviour, vol. 4, no. 8, pp. 856-865, 2020.

[12] P. Mecenas, R. Bastos, A. Vallinoto, and D. Normando, “Effects of temperature and humidity on the spread of covid-19: A systematic review.,” medRxiv, 2020.

[13] A. Björck, Numerical Methods for Least Squares Problems. Society for Industrial and Applied Mathematics, 1996.

[14] C. Lawson and R. Hanson, Solving Least Squares Problems. Society for Industrial and Applied Mathematics, Jan. 1995.

[15] R. Burden, D. Faries, and A. Burden, Numerical Analysis. Cengage Learning, 2014.

[16] M. Ruggiero and V. Lopes, Cálculo Numérico: Aspectos Teóricos e Computacionais. Pearson Universidades, 2000.

[17] D. Sperandio, J. T. Mendes, and L. H. Monken, Cálculo Numérico. Pearson Universidades, 2014.

[18] https://covid.saude.gov.br.

[19] https://www.ins.gov.co.

[20] https://coronavirus.jhu.edu/map.html.

[21] T. McMichael and et al., “Epidemiology of covid-19 in a long-term care facility in king county, Washington,” N. Engl. J. Med., vol. 382, pp. 2005-2011, Mar. 2020.

[22] D. Studdert and M. Hall, “Disease control, civil liberties, and mass testing - calibrating restrictions during the covid-19 pandemic,” N. Engl. J. Med., vol. 383, pp. 102-104, Apr. 2020.

[23] E. Cirilo, S. Petrovskii, and P. Romeiro, N.and Natti, “Investigation into the critical domain problem for the reaction-telegraph equation using advanced numerical algorithms,” Int. J. Appl. Comput. Math., vol. 5, no. 3, pp. 54-, 2019.

[24] E. Piccolomini and F. Zama, “Preliminary analysis of covid-19 spread in italy with an adaptive seird model.” https://arxiv.org/abs/2003.09909, March 2020.

[25] E. Piccolomini and F. Zama, “Monitoring italian covid-19 spread by a forced seird model,” PLOS ONE, vol. 15, pp. 1-17, March 2020.

